# Understanding Reach, Effectiveness, Adoption, Implementation, and Maintenance of home-based comprehensive sexual health care: a Realist Review

**DOI:** 10.1101/2023.11.02.23297983

**Authors:** Cornelia J.D. Goense, Thuan-Huong P. Doan, Eneyi E. Kpokiri, Ymke J. Evers, Claudia S. Estcourt, Rik Crutzen, Jeffrey D. Klausner, Weiming Tang, Paula Baraitser, Christian J.P.A. Hoebe, Nicole H.T.M. Dukers-Muijrers

## Abstract

**Background:** Testing for *human immunodeficiency virus* (HIV) and sexually transmitted infections (STI) is increasingly offered outside a clinic-based setting. Among key populations barriers to accessing testing and sexual health care may could be lowered due to home-based testing and care services. This review identifies which elements of home-based comprehensive sexual health care (home-based CSH) impacted which key populations, under which circumstances.

**Methods:** A realist review of studies focused on home-based CSH with at least self-sampling or self-testing HIV and additional sexual health care (e.g., treatment, counselling). PubMed, Embase, Cochrane Register of Controlled Trials, and PsycINFO databases were searched. Peer-reviewed quantitative and qualitative literature published between February 2012 and February 2023 was examined. The RE-AIM framework was used to systematically assess the (R) reach of key populations, (E) effectiveness of the intervention, and (AIM) effects on the adoption, implementation, and maintenance within routine sexual health care.

**Results:** Of 730 uniquely identified records, 92 were selected for extraction. Of these studies, 59% (54/92) reported actual interventions and 41% (38/92) described the acceptability and feasibility. Studies were mainly based in Europe or North America and were mostly targeted to MSM (59%; 55/92) (R). Overall, self-sampling or self-testing was highly acceptable across key populations. The effectiveness (E) of most studies was (expected) increased HIV testing. Adoption (A) of the home-based CSH was acceptable for care providers if linkage to care was available, even though a minority of studies reported adoption by care providers and implementation fidelity (I) of the intervention. In terms of maintenance (M), home-based CSH should be institutionalised and complementary to clinic-based sexual health care.

**Conclusions:** Five key findings were identified which may enhance implementation of home-based CSH. When providing the individual with a choice of testing, clear instructions, and tailored dissemination successful uptake of HIV testing may increase. For implementers perceived care and treatment benefits for clients may increase their willingness to implement home-based CSH. Therefore, home-based CSH may determine more accessible sexual health care and increased uptake of HIV testing among key populations.

## BACKGROUND

Sexually transmitted infections (STI) such as *Chlamydia trachomatis* (CT), *Neisseria gonorrhoeae* (NG), Hepatitis B (HBV), Hepatitis C (HCV), syphilis and human immunodeficiency virus (HIV) constitute a public health concern among key populations world-wide [1]. Key populations include populations vulnerable to HIV transmission such as men who have sex with men (MSM), sex workers, and transgender people. If STI or HIV are left undiagnosed and untreated, this could lead to serious adverse health consequences including acute illness, infertility and, long-term disability [2]. Regular testing followed by timely treatment is an effective strategy to improve clinical outcomes. Barriers to regular testing include social barriers such as expected HIV-related stigma and structural barriers such as costs or distance to the clinic [3, 4]. Home-based interventions, including self-(sampling) testing for HIV and STI, have gained urgency and popularity, especially during the last years of the COVID-19 pandemic [5]. Home-based interventions may lower commonly reported barriers and enhance personal autonomy thereby increasing accessibility and uptake of testing in key populations [6–8].

Home-based comprehensive sexual health care (hereafter: home-based CSH) includes either self-sampling or self-testing STI/HIV and additional sexual health care such as treatment or counselling. Self-sampling is when a person collects samples and sends them to a laboratory for testing, self-testing is when a person samples and interprets the test results themselves [9]. Self-sampled and clinician-collected samples provide comparable results for CT and NG testing, similar to the performances and usability of several HIV self-tests[10]. Therefore, the World Health Organisation (WHO) has recommended making self-sampling or self-testing STI and HIV available in addition to clinic-based sexual health services [8].

Earlier reviews about home-based CSH have mostly focused on its outcomes in terms of effectiveness and patient experiences of the care offered, with less attention to the implementation process in real-world settings [11–13]. The realist approach is especially relevant for understanding complex interventions, such as home-based and sexual health care [14]. Where systematic reviews usually aim to answer whether an intervention provides desired results, a realist review is particularly concerned with understanding why and how an intervention may or may not work, under which circumstances, and for which populations [15].

This realist review aimed to provide an overview of the mechanisms and contextual factors that determine what elements within home-based CSH impact which key populations under what circumstances. The reach, effectiveness, adoption, implementation and maintenance framework (RE-AIM) was used as structural guidance, which is a widely used framework to assess the impact of public health interventions [16]. This realist review used existing literature to identify working mechanisms and outcomes, which are influenced by contextual factors such as different settings and populations, thereby providing valuable information for key populations, care providers and policymakers.

## METHODS

We conducted a realist review, which required the construction and refinement of an initial programme theory. Figure 1 shows the initial programme theory that assessed context, mechanisms and outcomes. Outcomes were determined by reach(R), effectiveness(E), adoption(A), and implementation fidelity(I) elements of home-based CSH and how it affect the maintenance(M) of interventions [16]. We conducted the following iterative steps: 1) clarified research scope, 2) searched for relevant evidence, 3) appraised and extracted data, 4) synthesised evidence and 5) evaluated findings [14]. This review is reported following the publication standards realist and meta-narrative evidence Syntheses (RAMESES) guidelines for realist synthesis [17]. This study is pre-registered in PROSPERO (CRD42023397383).

**Figure 1.**
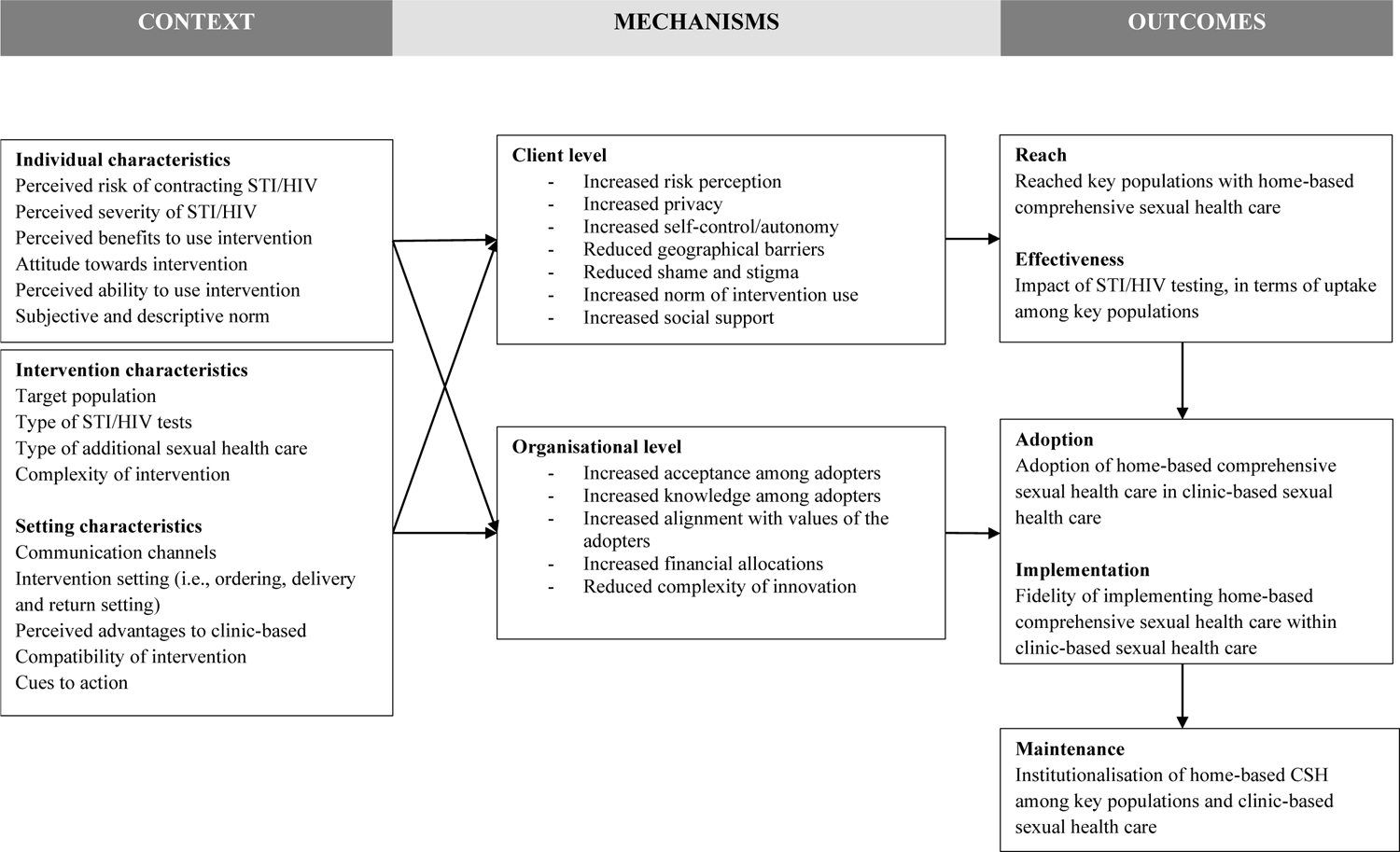
Initial programme theory: Theoretical framework of context, mechanisms and outcomes

### Inclusion criteria

Eligible interventions had a self-testing or self-sampling testing component. The intervention of home-based comprehensive sexual health care must consist of HIV testing alone or combined with other STI tests performed by the individual outside of a clinic location. In addition, the interventions should have other care components, such as sexual health counselling, partner notification or, linkage to treatment. Interventions could target any key population at elevated risk for HIV infection, such as men who have sex with men, sex workers or transgender people.

### Evidence identification

PubMed, Embase, PsycINFO and Cochrane Central Register of Controlled Trials (CENTRAL) were searched [18]. The initial search aimed to explore initial key theories in the several interventions and refine inclusion criteria. The initial search was conducted in February 2022, and analysis and selection of articles was completed in June 2022 and covered literature from the past 10 years [see Additional file 1]. We adjusted the search strategy according to different terminology used to refer to home-based CSH in studies (i.e., telehealth, self-managed testing, home-testing). The first author conducted the initial search and together with the second and third authors performed independent title, abstract and full-text screening. Literature was selected based on pre-specified inclusion and exclusion criteria [see Additional file 2]. The first three authors independently reviewed and analysed a random selection of approximately 200 to 300 unique records each, all authors approved of the final selection.

### Data extraction

Figure 2 shows that from a total of 730 unique records identified, and after exclusion of 620 records, 106 studies (17%) had been initially selected for review. The main reasons for exclusion were ‘intervention did not include HIV testing’ or ‘HIV testing is not self-sampling or self-testing’. Two reports were added to the selection using snowballing (i.e., referenced by included articles but not yet included in the search). An iterative selection excluded another 25 studies, mostly because they were systematic reviews or did not fit the research purpose. The final search added 11 studies from February 2022 to February 2023.

**Figure 2.**
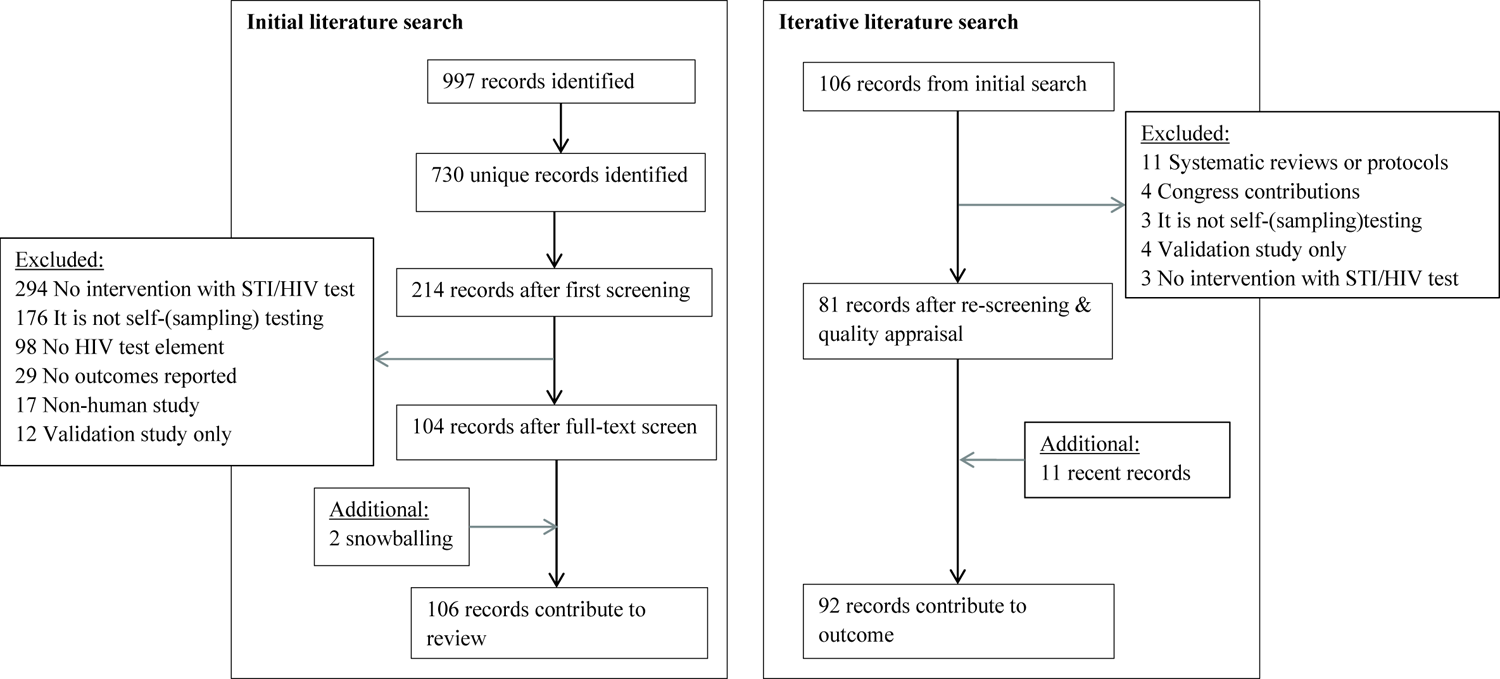
Flowchart of study search strategy through an iterative search process

### Quality appraisal

The quality of the literature was determined by judging to what extent the selected literature contributes to the outcome of the study questions. The selection was appraised by following the mixed methods appraisal tool (MMAT) and additional questions were assessed such as a) did the study describe constructs of the initial programme theory (Figure 2) b) were intervention components described, d) was the context of the intervention described and e) did the study describe the way the intervention affects outcomes (of RE-AIM) [19]. Studies were considered sufficient quality if clear research questions were reported, and the data allowed to address the research question. The first author conducted the quality appraisal which was randomly checked by the second and third author, inconsistencies were decided upon among the first three authors [see Additional file 3]. All authors agreed with the overall selection procedure. The data extraction process is mainly conducted by the first author, other authors verified the process and provided feedback. Results were presented using the RE-AIM framework. In addition, enhancing process factors were formulated based on evidence from all included studies and each included context (C), mechanisms (M) and outcomes (O) components.

## RESULTS

A total of 92 studies were included that examined interventions of home-based CSH with STI and HIV self-sampling or self-testing and additional sexual health care [see Additional file 4]. Most studies (59%; 54/92) described research with intervention components in randomized, non-randomized, quasi-experimental (qualitative and quantitative) evaluation studies. The other studies (41%; 38/92) solely described the acceptability or intention of key populations to potentially use an intervention (hereafter: acceptability studies). Overall, most studies were from Europe (n=30) or North America (n=28) and focused on the key populations MSM (n=55), sex workers (n=7) and transgender people (n=6). Based on the evidence from all 54 evaluation studies with intervention components tested in real-life settings, five statements were described. Results were enriched with information from 38 acceptability studies. Table 1 shows a summary of enhancing process factors. Process factors on the level of both the intervention and the implementation contribute to successfully implementing home-based CSH in practice and maintenance of its value for sexual health care of key populations in the long term.

**Table 1.**
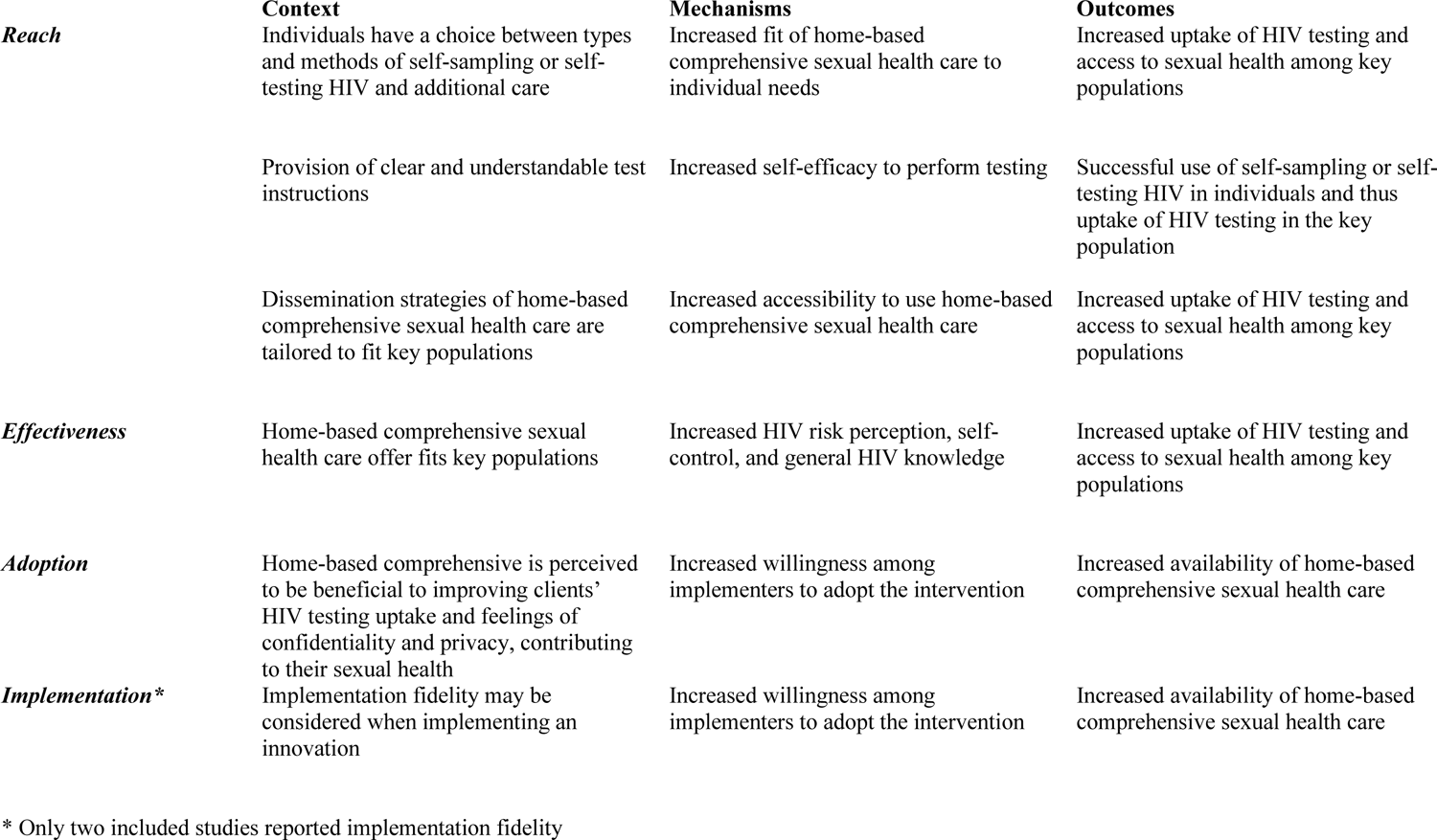
Key findings that enhance implementation of home-based comprehensive sexual health care.

### Reach

The type of self-sampling or self-testing is an important factor for the acceptance and use of home-based CSH. Most included studies assessed HIV self-testing (54%; 30/54) or self-sampling HIV testing (33%; 18/54) interventions, and 16 (11%) intervention studies were combined with self-sampling STI testing. In a study among the general population of the United Kingdom, self-testing was mostly preferred, and self-sampling testing when the ability to complete the test within the laboratory [20]. Samples required for HIV testing were either blood or saliva. Blood-based samples were more accurate for syphilis, HBV, HCV and HIV testing [21–23]. Blood was collected via finger-prick for dried-blood spot (DBS) or capillary collection (in a tube) or by a lancet in a vacuum system. The use of a finger-prick was acceptable to people in key populations across most interventions. It was feasible and could be done successfully [24, 25]. Reported challenges were mainly a result of insufficient knowledge of correct performance [26–28]. Blood collection with a vacuum system was highly acceptable and feasible for sampling among PrEP-using MSM. They were confident of performing this without supervision at home [29]. Saliva-based sampling was also experienced as acceptable and mostly easy to perform [27, 30–34]. In addition, saliva-based samples had a higher return rate than blood samples [23]. Most studies were restricted to results of a single type of testing, either blood-based or saliva-based. However, interventions that assessed both, recommended that both self-testing and self-sampling HIV testing should be available [24, 35].

Instructions to use the test kit were available before the actual testing and demonstrated by a care provider or study staff, or instructions were provided during testing with a video or written instructions. Several interventions did not specify the form in which instructions were provided. Clear instructions were reported to increase the usability of self-(sampling) testing [26, 34, 36, 37]. Complementary (photo or animated) images of the puncture site could support correct performance [24]. Studies among African populations highlighted the importance of instructions in local languages [34, 38, 39]. Interventions in younger key populations reported high acceptability of online or mobile application instructions [40–42] Various dissemination strategies were used to offer home-based CSH to key populations, and several venues were targeted. These include community venues, sex-on-premises, online services or existing (location-based) sexual health care. Only a few studies elaborated on the method to design dissemination methods. An example is the use of an intervention mapping framework to fit the communication messages, channels, and strategies to the needs of the key population [31, 43]. Other interventions only mentioned the communication modes (online or offline) [42, 44–46]. Online dissemination was mostly preferred by key populations when privacy concerns were related to clinic-based care [27, 35, 47, 48]. For key populations with inadequate access to digital technology, additional offline options (i.e., locations visited by key populations) should be available [49, 50]. Several studies demonstrated that online or peer-to-peer dissemination might be a promising strategy for reaching first-time testers [42, 46, 51–55]. For sex workers, self-tests were reportedly distributed through their workplaces (i.e., sex on premises, saunas, brothels), mobile units, and via online personal messages [41, 56, 57]. To recruit vulnerable key populations such as transgender people, online geotargeting was used. In addition, the costs of HIV self-(sampling) testing are mentioned as a barrier to testing uptake, and this is important to consider [58, 59].

### Effectiveness

The main reason organizations implement home-based CSH is to increase HIV testing in the people who need it [6–8]. The effectiveness of an accessible care offer could thus be identified by the numbers of test uptake, HIV diagnosis, linkage to sexual health care and prevention measures. Most studies demonstrated the identification of new HIV diagnoses. Overall, though HIV positivity rate varied between different contexts and populations, from 0.2% in a British study to 14.3% in a Chinese study among MSM [32, 60]. A substantial number of studies did not identify any new positive HIV results [20, 26, 30, 31, 33, 34, 40, 53, 61, 62]. Those studies were mainly based in Western continents (Europe, North America) and are focused on MSM. In studies that did not report positivity rates, the effectiveness of home-based care was attributed to the uptake of testing among at-risk and previously untested people and the return rate of test kits [51, 63–65]. Several interventions demonstrated high intention among populations to use home-based CSH again [33, 37, 40, 46, 66]. Home-based CSH may also affect other aspects of behavioural change of the individual, such as increased self-control, increased risk perception, and increased general knowledge of STI and HIV [33, 56, 62, 67]. Of 21 intervention studies that identified new HIV diagnoses, most of them also reported linkage to care such as confirmatory testing, linkage to treatment, post-test counselling, PrEP care, and partner notification. A few studies linked all new HIV infections successfully to care [59, 68]. Some studies reported referral, although it remained unknown whether clients had acted upon the referral to health care services [57, 69]. Other studies described loss to follow-up after positive HIV tests [45, 49, 60, 69]. Linkage to care in standard care is > 95%, and the linkage to care proportions of home-based care are comparable to outreach standards, i.e., within 75%-100% [42]. There were 13 studies that did not describe which additional sexual health options were offered (e.g., linkage to treatment, counselling, partner notification).

### Adoption

Here, seven studies are described that examined adoption by implementers and conditions for potential implementation in practice. Implementers are considered the individuals (i.e., care providers or counsellors) who enable access to home-based CSH or handle the patient management of people who are using the intervention. Implementation of home-based CSH is needed for the availability and, thereby the accessibility of the offer. Most studies described experienced or perceived facilitators and barriers to adopting home-based CSH [25, 31, 33, 43, 47, 70]. Overall, adoption was acceptable according to implementers as a result of facilitators such as increased access for clients to testing and increased confidentiality and privacy for clients. Implementers also mentioned the expected extended reach of vulnerable key populations and justification of clinical time (i.e., reduced or reallocated workload) [43, 70]. One of the concerns of adopting home-based CSH was expected missed opportunities for appropriate sexual health prevention information and interventions (e.g., vaccinations), that could have been given when consultations take place in person [27, 28, 31, 70]. Other concerns were regarding following up on people with positive results when testing outside a clinic[33, 71]. Expected pragmatic challenges were in data collection and reporting. Tailored programmes may be required for implementing home-based care, although that could result in complex data management [25, 72]. Implementing a secured digital platform for data may improve access. A few studies described their challenges with digital applications such as missing links between test kits and participants, or survey errors [25, 40]. Digital applications for home-based CSH were mainly created for use in high-income countries and may be less applicable to lower-income countries [72].

### Implementation

Implementation fidelity indicates the extent to which designed elements were used by implementers, as intended. Implementers, such as care providers, may have limited role tasks in carrying out the intervention since home-based CSH is designed for use by clients outside the clinic setting. Little information is available on the implementation fidelity of home-based interventions. One study demonstrated fidelity in the performance of the intervention. In one study, the performance fidelity of oral HIV self-testing was lower than expected since more than half of the participants had observed errors in their tests [73]. Another study described on an organisational level that a standardised script was recommended (i.e., a written version of all relevant elements) when introducing home-based CSH among care providers and their clients. Although the intervention was designed with these standardised elements, in practice tailoring was applied, which contributed to increasing the speed of processes [31]. Lastly, one study suggests simplification of intervention processes should contribute to implementation of home-based CSH, but without specifying the process simplification [59].

### Maintenance

The maintenance CSH assessed to what extent home-based CSH becomes part of routine sexual health care. Most studies suggested that home-based CSH may be offered complimentary to existing health care initiatives. For instance, a North American study evaluated the implementation of several oral HIV self-test interventions between 2018 and 2020, however, it did not examine its potential institutionalisation within existing sexual health care [25]. One study demonstrated that a self-sampling service among MSM increased awareness and intention to test, two years after implementation [74]. In most cases, the long-term impact of most home-based CSH has not yet been examined, since it has been implemented in recent years.

## DISCUSSION

Key populations are disproportionately affected by human immunodeficiency virus (HIV) and sexually transmitted infections (STI). Home-based comprehensive sexual health care (home-based CSH) can overcome barriers to accessing testing and care. This realist review assessed which elements of home-based CSH works for which key populations, and under which circumstances. A realist approach allowed for comparing key elements in a real-life context following the reach, effectiveness, adoption, implementation, and maintenance (RE-AIM) framework.

From a user perspective, enhancing elements were identified, such as the choice of either home-based or clinic-based HIV testing, clear instructions, and tailoring of dissemination, which may contribute to the uptake of HIV testing. Choice in the type of HIV testing should increase the fit of the offer to individual needs. A recent systematic review of randomised controlled trials demonstrated a contribution to the uptake of HIV testing when offering self-testing instead of standard-of-care testing [75]. Several studies indicated a higher uptake of HIV testing as a result of implementing home-based CSH [46, 61]. However, another systematic review demonstrated growing inequality, with vulnerable groups less represented among home-based CSH users. Home-based CSH may not be suitable when people need further clinical evaluation in case of poor sexual health and or digital literacy or for vulnerable people [23]. Therefore, home-based CSH should be offered in addition to clinic-based sexual health care in order for people to have a choice in the type of HIV testing and care they prefer [50]. Furthermore, an understandable and clear provision of test instructions could increase the self-efficacy of users. Previous studies identified that self-collected swabs and urine samples could be an alternative to swabs collected by a clinician [76]. Therefore, it is essential to offer clear instructions (i.e., written, illustrated or video) to ensure correctly taken samples [77]. A recent study among MSM and transgender people found users preferred video instructions when using an HIV self-test. In addition, inaccuracies in the interpretation of HIV testing results were reduced, when using visualized instructions[78]. Moreover, previous studies suggested that if dissemination strategies are tailored, home-based CSH will be more accessible to key populations [39]. Qualitative evaluation of a self-collection program assessed that the use of multiple communication channels is preferred [79]. In addition, a previous literature review of oral HIV self-testing emphasizes making the offer fitting and accessible for the needs of the key population in terms of language, sexual health literacy and culture [80].

From the implementers’ perspective, few insights on home-based CSH were demonstrated. Even though adoption by implementers may be essential to successfully implement and maintain an intervention. In a previous study, care providers found the distribution of home-based CSH highly acceptable. However, there was concern about which care provider or health care organisation should be responsible for managing clients when testing at home [81]. On an organisational level, the cost-effectiveness for finding new HIV infections is a facilitator to adopt home-based solutions [31, 41, 57]. However, at first, home-based CSH may have higher costs due to starting up processes required for implementation (e.g., collaboration with stakeholders, monitoring) [82]. Another concern is the capability of proper performance of HIV self-testing by clients [81, 83]. A complete care package with self-testing and additional web-based information or counselling may improve the acceptability of home-based CSH by care providers [84]. In addition, simplification of tests should contribute to the ability of clients to perform their home-based testing accurately. Further, self-sampling testing (i.e., when clients send their samples to a laboratory) might offer care providers more control over clients’ tests results and follow-up if necessary [20].

### Scientific implications

Terms such as telehealth, eHealth, and self-managed care, among other terminology, are used interchangeably to refer to different elements within home-based CSH. The WHO has called for clear and transparent definitions of digital health and self-care [85]. Earlier, the nomenclature was determined for self-testing and self-sampling testing [9]. To unravel universal success factors of home-based CSH, more comparative research and the reporting of processes in more detail, are warranted. Additionally, universal nomenclature in scientific articles may contribute to increased comparability and transparency of studies.

### Strengths and limitations

To our knowledge, this is the first realist review that aimed to examine enhancing elements of global home-based CSH. In addition, the realist approach seemed appropriate for assessing this complex intervention. However, a high number of studies had different definitions for home-based CSH and could therefore not be compared properly in such syntheses. Furthermore, a limited number of studies were included from implementation and maintenance perspectives. This could result in missing important conditions for implementation in the realist setting and the long-term impact of home-based CSH. As most included studies focused on MSM, information is lacking for other key populations, a common occurrence [75, 86, 87]. Therefore, future research should consider sampling key populations such as migrants, transgender people and sex workers.

## CONCLUSIONS

Home-based comprehensive sexual health care (home-based CSH) interventions have been implemented globally in recent years. This realist review aimed to identify which elements within home-based CSH works for which key populations under which circumstances. Five key elements for enhancing the reach, effectiveness, adoption and implementation of interventions were identified; choice of testing to fit individual needs, provision of clear instructions, tailored dissemination, increased individual determinants for successful uptake and, perceived care and treatment benefits for clients. Considering these elements within home-based CSH may result in more accessible sexual health care for key populations and increased uptake of HIV testing and care among these populations.

## Supporting information

Additional file 1, search strategy

Additional file 2, inclusion criteria

Additional file 3, quality appraisal

Additional file 4, included studies

## Abbreviations

CT: *Chlamydia Trachomatis,*

CSH: comprehensive sexual health care

DBS: dried blood spot

HBV: Hepatitis B

HCV: Hepatitis C

HIV: Human immunodeficiency virus

MSM: men who have sex with men

NG: *Neisseria Gonorrhoea*

RCT: randomized controlled trial

RE-AIM: reach, effectiveness, adoption, implementation and maintenance framework

STI: sexually transmitted infections

WHO: World Health Organisation

## Data Availability

Data sharing is not applicable since no datasets were generated or analysed during this study.

## Acknowledgements

First, we would like to thank Lucie Zeches for her cooperation in setting up this study and initiating the search strategy. We also want to thank Rianne de Wit for her contributions to the quality appraisal of the review.

## Availability of data and material

Data sharing is not applicable since no datasets were generated or analysed during this study.

## Authors contributions

All authors decided upon an initial research design. CG, T-HD, and EK searched and selected relevant literature. CG performed a synthesis of literature data, supervised by YE and ND. CG wrote the first draft of the paper; all authors approved the final draft before submission.

## Ethics approval and consent to participate

Not applicable.

## Consent for publication

Not applicable

## Additional files

**Additional file 1 - search strategy.pdf**

PDF

*Additional file 1. Initial search strategy and search strings*

The initial search strategy by database and search engine, with associated search strings

**Additional file 2 - inclusion.pdf**

PDF

*Additional file 2. Inclusion and exclusion criteria*

Inclusion and exclusion criteria which were used for the selection of studies.

**Additional file 3 - quality appraisal.xlsx**

XLSX

*Additional file 3. List of included studies and their quality appraisal*

Quality appraisal of all included studies by using the Mixed Methods Appraisal Tool (MMAT) and additional criteria.

**Additional file 4 – includedstudies.pdf**

PDF

*Additional file 4. Included studies that assessed home-based comprehensive sexual health care (n=92)*

All included studies with their study type, design, first author, year of publication, full title, study population and size (N) and continent of publication.

