## Additional file 1, search strategy for "Understanding Reach, Effectiveness, Adoption, Implementation, and Maintenance of home-based comprehensive sexual health care: a Realist Review"

*Additional file 1. Initial search strategy and search strings*

General search strategy

*Concept: HIV*

Keywords: HIV, human immunodeficiency virus
MeSH: "HIV"[Mesh]

Search string: (HIV OR human immunodeficiency virus OR “HIV” [Mesh])

*Concept: STI*

Keywords: sexually transmitted infection*, sexually transmitted disease, STI, STD, Gonorrhoeae, Gonorrhea, Chlamydia, Syphilis, Hepatitis B
MeSH: "Sexually Transmitted Diseases"[Mesh], "Chlamydia"[Mesh], "Gonorrhea"[Mesh], "Pre-Exposure Prophylaxis” [Mesh], “Syphilis” [Mesh], “Hepatitis B” [Mesh]

Search string: (“sexually transmitted infection*” OR gonorrhoea OR “Sexually Transmitted Diseases” [Mesh] OR “Chlamydia” [Mesh] OR “Gonorrhea’ [Mesh] OR “Pre-Exposure Prophylaxis” [Mesh] OR “Syphilis” [Mesh] OR “Hepatitis B” [Mesh])

*Concept: Test*

Keywords: test*, screen, diagnos*
MeSH: “HIV Testing” [Mesh], “Diagnosis” [Mesh]

Search string: (test* OR screen OR diagnos* OR “HIV testing” [Mesh] OR “Diagnosis” [Mesh])

*Concept: Method*

Keywords: self-collect*, self-sampl*, self-obtain*, “specimen collect*”, self-administ*, self-manage*, self-screen*, self-examin*
MeSH:-

Search string: (self-collect* OR self-sampl* OR self-obtain* OR "specimen collect*" OR self-administ* OR self-manage* OR self-screen* OR self-examin*)

*Concept: Location*

Keywords: internet, remote, digital, home, online, eHealth, mHealth, telehealth
MeSH: “Telemedicine” [Mesh], “Internet-Based Intervention” [Mesh], “Delivery of Health care” [Mesh]

Search string: (internet OR remote OR digital OR home* OR online OR “Telemedicine” [Mesh] OR “Internet-Based Intervention” [Mesh] OR “Delivery of Health Care” [Mesh] OR eHealth OR mHealth or telehealth)

Complete search strings

*PubMed → 546 results*

(“sexually transmitted infection*” OR STI OR STD OR gonorrhoea OR “Sexually Transmitted Diseases” [Mesh] OR “Chlamydia” [Mesh] OR “Gonorrhea” [Mesh] OR “Pre-Exposure Prophylaxis” [Mesh] OR “Syphilis” [Mesh] OR “Hepatitis B” [Mesh] OR HIV OR human immunodeficiency virus OR “HIV” [Mesh])
AND
(HIV OR human immunodeficiency virus OR “HIV” [Mesh])
AND
(test* OR screen OR diagnos* OR “HIV testing” [Mesh] OR “Diagnosis” [Mesh])
AND
(self-collect* OR self-sampl* OR self-obtain* OR “specimen collect*” OR self-administ* OR self-manage* OR self-screen* OR self-examin*)
AND
(internet OR remote OR digital OR home* OR online OR "Telemedicine"[Mesh] OR “Internet-Based Intervention” [Mesh] OR “Delivery of Health Care” [Mesh] OR ehealth OR mhealth or telehealth)
AND (y_10[Filter])

*Ovid search tool: Embase, MEDLINE, Cochrane Register of Controlled Trials → 375 results*
Filter: “Deduplicate; limit 2 to yr=2012-current”

(sexually transmitted infection* or STI or gonorrhoea or sexually transmitted diseas* or STD or chlamydia or gonorrhea or PrEP or syphilis or hepatitis* or HIV or human immunodeficiency virus) and (HIV or human immunodeficiency virus) and (test* or screen or diagnos*) and (self-collect* OR self-sampl* or self-obtain* or specimen collect or self-administ* or self-manag* or self-screen or self-examin*) and (internet* or remote or digital or home* or online* or Telemedicine or Internet-Based Intervention or Delivery of Health Care or eHealth or mHealth or telehealth)

*PsycInfo → 76 results*
Filter: “Publication Year: 2012-2022”

(sexually transmitted infection* or STI or gonorrhoea or sexually transmitted diseas* or STD or chlamydia or gonorrhea or PrEP or syphilis or hepatitis* or HIV or human immunodeficiency virus) and (HIV or human immunodeficiency virus) and (test* or screen or diagnos*) and (self-collect* OR self-sampl* or self-obtain* or specimen collect or self-administ* or self-manag* or self-screen or self-examin*) and (internet* or remote or digital or home* or online* or Telemedicine or telehealth or Internet-Based Intervention or Delivery of Health Care or eHealth or mHealth or telehealth)
