## Additional file 2, inclusion criteria for "Understanding Reach, Effectiveness, Adoption, Implementation, and Maintenance of home-based comprehensive sexual health care: a Realist Review"

*Additional file 2. Inclusion and exclusion criteria*

Inclusion criteria:

- Date of publication: up to 10 years ago
- Language: English
- Study design: ALL
- Population: primarily focused on key populations (e.g., young individuals, migrants, sex workers, MSM, transgender people, gender diverse people, bisexual and homosexual men, swingers, PWID, PLWHIV)
- Intervention: home-based comprehensive sexual health care with (online) self-sampling for HIV and/or STI testing and AT LEAST one of these care elements:
    - Results provision
    - Treatment
    - Partner notification
    - Health promotion/prevention
    - Retesting/repeat testing
- Studies should report AT LEAST one of these **outcomes** and/or **constructs*** (Following RE-AIM)

Exclusion criteria:

- Non-human studies
- Validation studies with only test related results (sensitivity, specificity)
- Intervention has no HIV testing component
    - Unless key population is HIV positive

*Table of constructs related to reach, effectiveness, adoption, implementation, and maintenance.

| RE-AIM | Answer to… | Signal words |
|---|---|---|
| **Reach** | "How do I reach the targeted population?" | Reach<br>Target<br>Number of …<br>Proportion of …<br>Represent |
| **Effectiveness** | "How do I know my intervention is effective?" | Efficacy<br>Impact<br>Effects /Affect<br>Effective(ness)<br>Influence<br>Outcome<br>Factors<br>Predictors |
| **Adoption** | "How do I develop organizational support to deliver my intervention?" | Adoption<br>Setting<br>Deliver<br>Support<br>Barriers<br>Facilitators |

| | | |
|---|---|---|
| **Implementation** | "How do I ensure the intervention is delivered properly?" | Implementation<br>Fidelity<br>Protocols |
| **Maintenance** | "How do I incorporate the intervention so that it is delivered over the long term?" | Maintenance<br>Policy<br>Incorporation<br>Long term<br>Follow-up |
