## Additional file 4, included studies for "Understanding Reach, Effectiveness, Adoption, Implementation, and Maintenance of home-based comprehensive sexual health care: a Realist Review"

*Additional file 4. Included studies that assessed home-based comprehensive sexual health care (n=92)*

| Type | Study design | First author | Year | Full title | N | Target population | Continent |
|---|---|---|---|---|---|---|---|
| Intervention | Non-randomized studies | Agusti | 2020 | Outreach HIV testing using oral fluid and online consultation of the results: Pilot intervention in Catalonia | 834 | MSM, (migrant)SW, TGW | Europe |
| Intervention | Mixed methods studies | Ahmed-Little | 2016 | Attitudes towards HIV testing via home-sampling kits ordered online (RUClear pilots 2011-12) | 3062 | General population (higher risk) | Europe |
| Intervention | Randomized controlled trial | Asiimwe | 2014 | Accuracy of un-supervised versus provider-supervised self-administered HIV testing in Uganda: A randomized implementation trial | 246 | Fishing community | Africa |
| Acceptability | Quantitative descriptive studies | Babatunde | 2022 | Assessment of knowledge and acceptability of HIV self-testing among students of selected universities in southwest Nigeria: an online cross-sectional study | 155 | Undergraduate students | Africa |
| Intervention | Non-randomized studies | Baraitser | 2019 | Using a Theory-based SMS/VM Intervention to Improve Sexual and Reproductive Health of Female Entertainment Workers in Cambodia | 1466 | General population | Europe |
| Intervention | Non-randomized studies | Belza | 2014 | Low knowledge and anecdotal use of unauthorized online HIV self-test kits among attendees at a street-based HIV rapid testing programme in Spain | 3340 | MSM and MSW | Europe |
| Intervention | Randomized controlled trial | Biello | 2021 | HIV self-testing and STI self-collection via mobile apps: experiences from two pilot randomized controlled trials of young men who have sex with men | QNT 80<br>QLT 37 | (Y)MSM | North America |
| Intervention | Non-randomized studies | Cannon | 2022 | Feasibility of a novel self-collection method for blood samples and its acceptability for future home-based PrEP monitoring | 141 | People who use PrEP[&] | North America |
| Intervention | Mixed methods studies | Carballo-Diéguez | 2012 | Willingness to participate in HIV therapeutic vaccine trials among HIV-infected patients on ART in China | 57 | MSM | North America |

| | | | | | | | |
|---|---|---|---|---|---|---|---|
| Intervention | Non-randomized studies | Catania | 2020 | Oral HIV Self-Implemented Testing: Performance Fidelity Among African American MSM | 178 | (AA) MSM | North America |
| Intervention | Mixed methods studies | Chasco | 2021 | Mixed-Methods Evaluation of the Incorporation of Home Specimen Self-Collection Kits for Laboratory Testing in a Telehealth Program for HIV Pre-exposure Prophylaxis | QNT 77<br>QLT 21 | People who use PrEP[&] | North America |
| Intervention | Non-randomized studies | Chiu | 2016 | Correlates of requesting home HIV self-testing kits on online social networks among African-American and Latino men who have sex with men | AA/L 76<br>MSM 36 | (AA/Latino) MSM | North America |
| Acceptability | Quantitative descriptive studies | Clarke | 2022 | Assessment of online self-testing and self-sampling service providers for sexually transmitted infections against national standards in the UK in 2020 | 31 | General population | Europe |
| Acceptability | Quantitative descriptive studies | Cushman | 2019 | Attitudes and preferences regarding the use of rapid self-testing for sexually transmitted infections and HIV in San Diego area men who have sex with men | 109 | MSM | North America |
| Acceptability | Qualitative study | Daniels | 2018 | Getting HIV self-test kits into the home for young African American MSM in Los Angeles: A qualitative report | 21 | (YAA) MSM | North America |
| Acceptability | Quantitative descriptive studies | d'Elbée | 2018 | Preferences for linkage to HIV care services following a reactive self-test: discrete choice experiments in Malawi and Zambia | 941 | (Black African) General population | Africa |
| Acceptability | Qualitative study | Dodds | 2018 | Acceptability of HIV self-sampling kits (TINY vial) among people of black African ethnicity in the UK: a qualitative study | FGD=12<br>IDI= 9 | People with African ethnicity | Europe |
| Intervention | Non-randomized studies | Driver | 2017 | HIV testing within the African community using home-based self collection of oral samples | 100 | African community | Australia |
| Acceptability | Quantitative descriptive studies | Dulai | 2021 | Awareness of and intention to use an online sexually transmitted and blood-borne infection testing service among gay and | 1272 | (GB)MSM | North America |

| | | | | | | | |
|---|---|---|---|---|---|---|---|
| | | | | bisexual men in British Columbia, two years after implementation | | | |
| Acceptability | Qualitative study | Eaton | 2018 | Do young black men who have sex with men in the deep south prefer traditional over alternative STI testing? | 36 | (YB)MSM | North America |
| Intervention | Non-randomized studies | Elliot | 2016 | Identifying undiagnosed HIV in men who have sex with men (MSM) by offering HIV home sampling via online gay social media: A service evaluation | 17361 | MSM, female sex workers(FSW), young women | Europe |
| Intervention | Non-randomized studies | Fisher | 2015 | Home sampling for sexually transmitted infections and HIV in men who have sex with men: A prospective observational study | 574 | MSM | Europe |
| Acceptability | Mixed methods studies | Flowers | 2017 | Preparedness for use of the rapid result HIV self-test by gay men and other men who have sex with men (MSM): a mixed methods exploratory study among MSM and those involved in HIV prevention and care | QNT 999 QLT 55 | MSM | Europe |
| Intervention | Mixed methods studies | Frasca | 2014 | Attitude and behavior changes among gay and bisexual men after use of rapid home HIV tests to screen sexual partners | 27 | (GB)MSM | North America |
| Acceptability | Qualitative study | Freeman | 2018 | Perceptions of HIV Self-Testing Among Men Who Have Sex With Men in the United States: A Qualitative Analysis | 46 | MSM | North America |
| Intervention | Mixed methods studies | Giguere | 2020 | Use of self-management tools in an online support service - Who, what, how? | 279 | TGSW | North America |
| Acceptability | Quantitative descriptive studies | Goense | 2022 | Intention to use and acceptability of home-based sexual health care among men who have sex with men who previously attended clinic-based sexual health care | 154 | (clinic visiting) MSM | Europe |
| Intervention | Non-randomized studies | Grov | 2016 | Recruiting A US national sample of HIV-negative gay and bisexual men to complete at-home self-administered HIV/STI testing and surveys: Challenges and opportunities | 5010 | (GB)MSM | North America |

| | | | | | | | |
|---|---|---|---|---|---|---|---|
| Intervention | Non-randomized studies | Grov | 2019 | Recruiting vulnerable populations to participate in HIV prevention research: findings from the Together 5000 cohort study | 1071 | At-risk men, TGW, TGM | North America |
| Acceptability | Quantitative descriptive studies | Han | 2014 | HIV self-testing among online MSM in China: implications for expanding HIV testing among key populations | 1342 | MSM | Asia |
| Acceptability | Quantitative descriptive studies | Hoyos | 2016 | Knowledge, actual and potential use of HIV self-sampling testing kits among MSM recruited in eight European countries | 8226 | MSM | Europe |
| Intervention | Non-randomized studies | Hoyos | 2013 | Never tested for HIV in Latin-American migrants and Spaniards: prevalence and perceived barriers | 5920 | Migrants | Europe |
| Acceptability | Quantitative descriptive studies | Iliyasu | 2020 | Acceptability and correlates of HIV self-testing among university students in northern Nigeria | 399 | University students | Africa |
| Intervention | Non-randomized studies | Jin | 2019 | An Internet-Based Self-Testing Model (Easy Test): Cross-Sectional Survey Targeting Men Who Have Sex With Men Who Never Tested for HIV in 14 Provinces of China | 879 | MSM | Asia |
| Intervention | Mixed methods studies | John | 2019 | Gay and bisexual men's experiences using self-testing kits for HIV and rectal and urethral bacterial sexually transmitted infections: Lessons learned from a study with home-based testing | QNT 1070<br>QLT 11 | MSM | North America |
| Acceptability | Mixed methods studies | Kaneko | 2022 | Increasing access to HIV testing for men who have sex with men in Japan using digital vending machine technology | QNT 222<br>QLT 54 | MSM | Asia |
| Acceptability | Qualitative study | Kelvin | 2020 | Choice in HIV testing: the acceptability and anticipated use of a self-administered at-home oral HIV test among South Africans | 20 | General population | Africa |
| Intervention | Randomized controlled trials | Kelvin | 2018 | Offering self-administered oral HIV testing to truck drivers in Kenya to increase testing: a randomized controlled trial | 305 | (Black African) Truck drivers | Africa |
| Intervention | Qualitative study | Knight | 2019 | Qualitative analysis of the experiences of gay, bisexual and other men who have sex with | 37 | MSM | North America |

| | | | | | | | |
|---|---|---|---|---|---|---|---|
| | | | | men who use GetCheckedOnline.com: a comprehensive internet-based diagnostic service for HIV and other STIs | | | |
| Acceptability | Quantitative descriptive studies | Lau | 2020 | Prevalence of and factors associated with behavioral intention to take up home-based HIV self-testing among male clients of female sex workers in China - an application of the Theory of Planned Behavior | 303 | (M)C(F)SW | Asia |
| Intervention | Non-randomized studies | Lebina | 2019 | Influence of CYP2B6 and ABCB1 SNPs on nevirapine plasma concentrations in Burundese HIV-positive patients using dried sample spot devices | 2061 | Men, young women and sex workers (SW) | Africa |
| Acceptability | Quantitative descriptive studies | Leenen | 2022 | HIV testing and associated factors in men who have sex with men by urbanization-level: A cross-sectional study in the Netherlands | 3815 | MSM | Europe |
| Acceptability | Mixed methods studies | Leenen | 2021 | Systematic Development of an Intervention to Promote Self-Sampling for HIV and Sexually Transmitted Infections for Men Who Have Sex With Men: An Intervention Mapping Approach | 110 | MSM | Europe |
| Intervention | Non-randomized studies | Lippman | 2018 | High Acceptability and Increased HIV-Testing Frequency After Introduction of HIV Self-Testing and Network Distribution Among South African MSM | 127 | MSM | Africa |
| Intervention | Mixed methods studies | Loos | 2016 | Acceptability of a Community-Based Outreach HIV-Testing Intervention Using Oral Fluid Collection Devices and Web-Based HIV Test Result Collection Among Sub-Saharan African Migrants: A Mixed-Method Study | QNT 780<br>QLT 10 | (African) Migrants | Europe |
| Intervention | Non-randomized studies | MacGowan | 2018 | Pilot evaluation of the ability of men who have sex with men to self-administer rapid HIV tests prepare dried blood spot cards and interpret test results Atlanta Georgia 2013 | 22 | MSM | North America |

| | | | | | | | |
|---|---|---|---|---|---|---|---|
| Intervention | Non-randomized studies | Maksut | 2016 | A Test of Concept Study of At-Home, Self-Administered HIV Testing With Web-Based Peer Counseling Via Video Chat for Men Who Have Sex With Men | 20 | (AA/Black)MSM | North America |
| Intervention | Non-randomized studies | Manavi | 2017 | Observational study of factors associated with return of home sampling kits for sexually transmitted infections requested online in the UK | 3099 | General population | Europe |
| Acceptability | Quantitative descriptive studies | Maté | 2020 | Potential of HIV self-sampling to increase testing frequency among gay, bisexual, and other men who have sex with men, and the role of online result communication: Online cross-sectional study | 5019 | (GB)MSM | Europe |
| Intervention | Non-randomized studies | Mbopi-Kéou | 2020 | Launching HIV self-testing in a multicultural setting in Cameroon | 322 | General population | Africa |
| Acceptability | Quantitative descriptive studies | Miners | 2019 | Preferences for HIV testing services among men who have sex with men in the UK: A discrete choice experiment | 620 | MSM | Europe |
| Acceptability | Quantitative descriptive studies | Mokgatle | 2017 | High Acceptability of HIV Self-Testing among Technical Vocational Education and Training College Students in Gauteng and Northwest Province: What Are the Implications for the Scale Up in South Africa? | 3662 | College students (TVET) | Africa |
| Intervention | Randomized controlled trial | Mulubwa | 2019 | Community based distribution of oral HIV self-testing kits in Zambia: a cluster-randomised trial nested in four HPTN 071 (PopART) intervention communities | HIVST 13267<br>NONST 13706 | General population | Africa |
| Intervention | Non-randomized studies | Nash | 2021 | Acceptability and usability of HIV self-tests in two European countries: findings from surveys of clients at non-governmental organisations in Lithuania and Italy | LT 138<br>IT 28 | General population | Europe |
| Acceptability | Qualitative study | Nnko | 2020 | Female sex workers perspectives and concerns regarding HIV self-testing: an exploratory study in Tanzania | 227 | Female sexworkers (FSW) | Africa |

| | | | | | | | |
|---|---|---|---|---|---|---|---|
| Intervention | Non-randomized studies | Norelli | 2021 | Scaling Up CareKit: Lessons Learned from Expansion of a Centralized Home HIV and Sexually Transmitted Infection Testing Program | 775 | MSM | North America |
| Acceptability | Quantitative descriptive studies | O'Donnell | 2019 | Inequalities in HIV testing uptake and needs among men who have sex with men living in Ireland: findings from an internet survey | 2770 | MSM | Europe |
| Acceptability | Qualitative study | Pérez | 2016 | I Know that I Do Have HIV but Nobody Saw Me': Oral HIV Self-Testing in an Informal Settlement in South Africa | 11 | General population | Africa |
| Intervention | Non-randomized studies | Phanuphak | 2020 | Linkages to HIV confirmatory testing and antiretroviral therapy after online, supervised, HIV self-testing among Thai men who have sex with men and transgender women | MSM 465<br>TGW 99 | MSM and TGW | Asia |
| Intervention | Non-randomized studies | Phanuphak | 2018 | Where do young men want to access STI screening? A stratified random probability sample survey of young men in Great Britain | 571 | MSM, TGW | Asia |
| Intervention | Qualitative study | Pierre | 2020 | A qualitative study on oral-fluid-based HIV self-testing experiences among men in Kigali, Rwanda | 21 | Men | Africa |
| Intervention | Non-randomized studies | Pintye | 2019 | Acceptability and outcomes of distributing HIV self-tests for male partner testing in Kenyan maternal and child health and family planning clinics | 3620 | Mothers (Women) | Africa |
| Intervention | Non-randomized studies | Platteau | 2015 | Symptom profile and technology use of people living with HIV who access services at a community-based organization in the Deep South | 1071 | MSM | Europe |
| Acceptability | Quantitative descriptive studies | Qin | 2017 | Benefits and Potential Harms of Human Immunodeficiency Virus Self-Testing Among Men Who Have Sex With Men in China: An Implementation Perspective | 1189 | MSM | Asia |
| Intervention | Non-randomized studies | Radebe | 2020 | HIV self-screening distribution preferences and experiences among men who have sex | 127 | MSM | Africa |

| | | | | | | | |
|---|---|---|---|---|---|---|---|
| | | | | with men in Mpumalanga Province: Informing policy for South Africa | | | |
| Intervention | Qualitative study | Raffe | 2020 | HIV self-tests for men who have sex with men, accessed via a digital vending machine: a qualitative study of acceptability | 23 | MSM | Europe |
| Intervention | Non-randomized studies | Rahib | 2022 | Online self-sampling kits for human immunodeficiency virus and other sexually transmitted infections: Feasibility, positivity rates, and factors associated with infections in France | 1948 | MSM | Europe |
| Intervention | Non-randomized studies | Rahib | 2021 | Online self-sampling kits to screen multipartner MSM for HIV and other STIs: Participant characteristics and factors associated with kit use in the first 3 months of the MemoDepistages programme, France, 2018 | 7158 | MSM | Europe |
| Intervention | Non-randomized studies | Ricca | 2016 | Factors Associated with Returning At-Home Specimen Collection Kits for HIV Testing among Internet-Using Men Who Have Sex with Men | 896 | MSM | North America |
| Acceptability | Quantitative descriptive studies | Robinson | 2017 | Preferences for Home-Based HIV Testing Among Heterosexuals at Increased Risk for HIV/AIDS: New Orleans, Louisiana, 2013 | 464 | At-risk heterosexuals | North America |
| Intervention | Non-randomized studies | Rosenthal | 2023 | Geospatial Prioritization to Reach Hispanic or Latino and Other Priority Populations Through HIV Home Testing Services | 2554 | Hispanic/Latino MSM and TG/GNC people who have sex with men | North America |
| Acceptability | Qualitative study | Ryu | 2023 | Disruptions of sexually transmitted and blood borne infections testing services during the COVID-19 pandemic: accounts of service providers in Ontario, Canada | 18 | Service providers | North America |
| Acceptability | Quantitative descriptive studies | Saunders | 2012 | Where do young men want to access STI screening? A stratified random probability sample survey of young men in Great Britain | 411 | Young men | Europe |

| | | | | | | | |
|---|---|---|---|---|---|---|---|
| Intervention | Mixed methods studies | Seguin | 2018 | Self-testing is a feasible and acceptable option for identifying extragenital gonorrhea (GC) and Chlamydia (CT) infections in HIV-Infected People | QNT 119<br>QLT 21 | Black Africans | Europe |
| Acceptability | Quantitative descriptive studies | Sharma | 2014 | Acceptability and intended usage preferences for six HIV testing options among internet-using men who have sex with men | 973 | MSM | North America |
| Intervention | Qualitative study | Sharma | 2022 | Perceptions and Experiences of Returning Self-collected Specimens for HIV, Bacterial STI and Potential PrEP Adherence Testing among Sexual Minority Men in the United States | 25 | (GB)MSM | North America |
| Intervention | Non-randomized studies | Sharma | 2022 | Acceptability and Feasibility of Self-Collecting Biological Specimens for HIV, Sexually Transmitted Infection, and Adherence Testing Among High-Risk Populations (Project Caboodle!): Protocol for an Exploratory Mixed-Methods Study | 100 | (GB)MSM | North America |
| Acceptability | Quantitative descriptive studies | Shrestha | 2022 | Mobile Health Technology Use and the Acceptability of an mHealth Platform for HIV Prevention Among Men Who Have Sex With Menin Malaysia: Cross-sectional Respondent-Driven Sampling Survey | 376 | MSM | Asia |
| Acceptability | Quantitative descriptive studies | Syred | 2019 | Choose to test: Self-selected testing for sexually transmitted infections within an online service | 6253 | General population | Europe |
| Acceptability | Qualitative study | Tobin | 2018 | Acceptability and feasibility of a Peer Mentor program to train young Black men who have sex with men to promote HIV and STI home-testing to their social network members | 15 | (YB)MSM | North America |
| Acceptability | Quantitative descriptive studies | Tonen-Wolyec | 2019 | Acceptability of HIV self-testing in African students: a cross-sectional survey in the Democratic Republic of Congo | 1012 | Students | Africa |
| Intervention | Non-randomized studies | Tonen-Wolyec | 2019 | Acceptability, feasibility, and individual preferences of blood-based HIV self-testing in | 628 | Adolescents | Africa |

| | | | | | | | |
|---|---|---|---|---|---|---|---|
| | | | | a population-based sample of adolescents in Kisangani, Democratic Republic of the Congo | | | |
| Acceptability | Non-randomized studies | Turner | 2018 | Web-based data collection for older adults living with HIV in a clinical research setting: pilot observational study | 5632 | General population | Europe |
| Intervention | Non-randomized studies | Van Loo | 2017 | Self-collected genital swabs compared with cervicovaginal lavage for measuring HIV-1 and HSV-2 and the effect of acyclovir on viral shedding | 217 | MSM | Europe |
| Acceptability | Mixed methods studies | Vera | 2019 | Acceptability and feasibility of using digital vending machines to deliver HIV self-tests to men who have sex with men | QNT 232<br>QLT 10 | MSM | Europe |
| Intervention | Non-randomized studies | Volk | 2016 | Acceptability and feasibility of HIV self-testing among men who have sex with men in Peru and Brazil | 103 | MSM | South America |
| Intervention | Non-randomized studies | Wong | 2022 | Regular Testing of HIV and Sexually Transmitted Infections With Self-Collected Samples From Multiple Anatomic Sites to Monitor Sexual Health in Men Who Have Sex With Men: Longitudinal Study | 204 | MSM | Asia |
| Intervention | Non-randomized studies | Wulandari | 2020 | Usability of a consumer health informatics tool following completion of a clinical trial: Focus group study | 292 | CSW (MWPS) | Asia |
| Intervention | Non-randomized studies | Xia | 2018 | Feasibility of an internet-based HIV testing service: anonymous urine collection from men who have sex with men | 1977 | MSM | Asia |
| Intervention | Randomized controlled trials | Xiao | 2020 | Sexual network distribution of HIV self-testing kits: Findings from the process evaluation of an intervention for men who have sex with men in China | 177 | MSM | Asia |
| Acceptability | Quantitative descriptive studies | Zhang | 2022 | The Impacts of the COVID-19 Pandemic on HIV Testing Utilization among Men Who Have Sex with Men in China: Cross-sectional Online Survey | 595 | (GB)MSM | Asia |

| | | | | | | | |
|---|---|---|---|---|---|---|---|
| Intervention | Non-randomized studies | Zhong | 2017 | Acceptability and feasibility of a social entrepreneurship testing model to promote HIV self-testing and linkage to care among men who have sex with men | 380 | MSM | Asia |

[&] PrEP indications as stated by CDC PrEP guidelines https://www.cdc.gov/hiv/clinicians/prevention/prep.html
